# Comparing the Natural Decline of Carbon Monoxide (CO) Levels Using Four Breath CO Monitors Among Hospitalized Daily Tobacco Smokers: Implications for Determining Abstinence in Smoking Cessation Clinical Trials

**DOI:** 10.64898/2026.11.20.26368591

**Authors:** William P. Wagner, Brionna Smith, Dustin P. Long, Stephen J. Thomas, Michael Businelle, Karen L. Gamble, Karen L. Cropsey

## Abstract

Biochemical verification of smoking outcomes is standard for determining the efficacy of treatments in smoking cessation clinical trials. The most common, cost-effective, and non-invasive biochemical verification method is breath carbon monoxide (CO) monitoring. No studies have examined the natural course of CO reduction in forced abstinence settings over multiple days with multiple devices to determine optimal cutoffs. This project compared the four most commonly used CO monitors (Vitalograph BreathCO, piCO+, iCOquit [corded and Bluetooth]) across 44 hours of monitoring on an inpatient hospital unit for 26 participants who reported daily cigarette smoking. Linear mixed models were used to fit smooth curves and control within subject correlations. Receiver operator characteristic (ROC) analysis was performed using logistic regression to examine performance of different cut-offs for CO for each device using sensitivity, specificity, Youden’s J, area under the curve (AUC), and efficiency. By the end of monitoring (>40 hours), 100% of participants had a ≤3 ppm on Vitalograph BreathCO. Using the Vitalograph BreathCO as the gold standard, both the bluetooth and corded iCOquits had an optimal cutoff of ≤3 ppm, while the piCO+ had an optimal cutoff of ≤5 ppm. CO monitors demonstrated different cutoffs and should not be used interchangeably. Cutoffs higher than 5 ppm should not be used to indicate abstinence, regardless of CO device.

## 1 INTRODUCTION

Combustible cigarette smoking remains the leading cause of death and disability and is the only legally sold product in the United States that results in death when used as intended for half of all consumers (Doll et al., 2004). About 9.9% of adults smoke cigarettes (Agaku, 2026; VanFrank et al., 2024), with smoking rates decreasing significantly over the past 60 years(Arrazola, 2025). The decline in smoking rates is attributed to changing policies (e.g., cigarette taxes, smoke-free public spaces), fewer teens initiating cigarette smoking, the public health success in messaging the dangers of tobacco use, and the availability of better treatments for smoking cessation (Cummings, 2016).

Determining smoking abstinence during a clinical trial is critical for evaluating the effectiveness of new cessation interventions (Benowitz et al., 2020a). The gold standard for confirming self-reported abstinence is biochemical verification, commonly measured from urinary or salivary cotinine or breath carbon monoxide (CO) (Benowitz et al., 2020a). Breath CO has several advantages over cotinine. Measuring it is non-invasive and less expensive, and CO readings can be used in trials that include nicotine replacement products (Benowitz et al., 2020b). More recently, with remote or decentralized clinical trials, breath CO devices that connect to data collection apps via Bluetooth (e.g., iCOquit) can be mailed to participants to verify smoking cessation, with CO readings automatically transmitted to investigators (Dahne et al., 2024; Thrul et al., 2023; Tuck et al., 2021).

CO cutoffs indicating abstinence were initially set at 8 to 10 parts per million (ppm), regardless of the model of CO monitor used in cessation trials (SRNT Subcommittee on Biochemical Verification, 2002). However, more recent studies show that cutoffs of 3 to 6 ppm may be more accurate for correctly classifying abstinence (Benowitz et al., 2020b; Cropsey et al.,

2006, 2014; Ramani et al., 2023; Tonkin et al., 2023). These studies tested breath CO’s reliability by comparing readings between people who smoke and non-smokers (Cropsey et al., 2006; Ramani et al., 2023), between breath CO and urine cotinine (Cropsey et al., 2014), and between different CO monitors (Tonkin et al., 2023; Tuck et al., 2021). To our knowledge, no study has examined the natural course of breath CO dissipation in a controlled environment (e.g., hospital setting) where smoking is not allowed. This is important as prior studies relied on participants’ self-reports of recent tobacco use or abstinence, which may have been reported inaccurately (Cropsey et al., 2014; Tonkin et al., 2023). Thus, the present study aimed to assess the variability of breath CO readings between different CO devices by measuring the natural decline of CO during a monitored, multi-day period of smoking abstinence. We compared the four most commonly used breath CO monitors in research studies: the Vitalograph BreathCO, the Bedfont piCO+, the Bedfont corded iCOquit Smokerlyzer and the more recent Bluetooth iCOquit Smokerlyzer.

## 2 METHODS

### 2.1 Participants

Participants were recruited from the Birmingham, Alabama community as part of a larger study examining racial differences in sleep and smoking behavior (R01DA046096). Participants were eligible for the original study if they were 18 years or older, identified as either Black or White, and smoked five or more cigarettes per day. Exclusion criteria included a serious medical or psychiatric conditions; a sleep disorder other than insomnia; severe sleep apnea, as measured by an apnea-hypopnea index; any current use of stimulants, opioids, benzodiazepines, sedatives, or hypnotics; a cannabis or alcohol use disorder; a positive pregnancy test; a self-report of current breastfeeding; or current employment as a night shift worker. All participants provided written informed consent to participate, and study procedures were approved by the University of Alabama at Birmingham IRB (IRB-300003205) and registered on ClinicalTrials.gov (NCT03968900).

### 2.2 Study Procedures

After providing informed consent, smoking status was confirmed with a urine cotinine test and breath CO ≥8 ppm using the Vitalograph BreathCO. Abstinence from stimulants, opioids, benzodiazepines, sedatives, and hypnotics was also confirmed with a urine test. Then participants completed surveys about their smoking behavior, dependence, cravings, withdrawal, and sleep behavior.

Participants completed two consecutive nights of sleep observation where they were randomized to receive two nights of sleep extension (10-hour sleep opportunity each night) or sleep restriction (4-hour sleep opportunity each night). The start time of each participant’s sleep opportunity was their self-reported bedtime. Participants received a two-week washout period before returning to perform the alternate sleep condition.

Participants were prompted to smoke a cigarette between 5:00 PM and 6:00 PM prior to entering the hospital on the first evening of sleep observation. At 6:00 PM, breath CO was measured using each of the four CO devices: corded iCOquit, Bluetooth iCOquit, Vitalograph BreathCO, and piCO+. Participants stayed on the hospital unit and were not allowed to smoke cigarettes or use nicotine products (e.g., nicotine replacement) for the 44-hour protocol. Outside of sleep opportunity windows, participants’ breath CO was measured every two hours using all four devices. Thus, breath CO was measured at 13 timepoints during the extended sleep condition and up to 21 timepoints for the restricted sleep condition. The measurement order of the CO devices was counterbalanced across timepoints.

### 2.3 Data Analyses

Average CO levels were calculated across time for all four devices to determine overall raw time trends. To examine CO differences over time between devices, linear mixed models were used to fit smooth curves and control for within subject correlations. Restricted cubic splines (RCS) were employed for smoothing using recommended default knot sets (Hastie et al., 2009). Using Vitalograph CO data, similar models examined sleep restriction condition, sex, and race differences. Differences in RCS curves were tested with RCS by factor interaction terms in the model. Additionally, receiver operator characteristic (ROC) analysis was performed using logistic regression to examine performance of different cut-offs for CO for each device using sensitivity, specificity, Youden’s J, area under the curve (AUC), and efficiency assuming that Vitalograph CO ≤3 ppm indicated having CO levels similar to non-smokers (Cropsey et al., 2006). For both Youden’s J and efficiency, larger values (i.e. closer to 1) indicate improved performance of the prediction. Youden’s J is calculated as *Sensitivity* + *Specificity* − 1.

Efficiency, which factors in the rate of “true negative” and “true positive” readings, is *Sensitivity* * *Prevalence* + *Specificity* * (1 − *Prevalence*), where Prevalence is the overall percentage of participants that reached Vitalograph CO ≤3 ppm. Similarly, time to reach this CO value was calculated, and the cumulative incidence function was estimated using Kaplan-Meier methods. All analyses were performed in SAS 9.4.

## 3 RESULTS

A total of 26 participants were included in the study. Participants had an average age of 47 years, were 50% Black (AA), and were 58% female. See Table 1 for additional participant characteristics. **Error! Reference source not found.Error! Reference source not found**. illustrates the raw CO averages over time for all four devices and shows that all but the corded Bedfont iCOquit monitors have similar values across time. The RCS model for these same values is shown in **Error! Reference source not found**., panels A-D. Thus, **Error! Reference source not found**.A is a smooth predicted version of **Error! Reference source not found**..

**Table 1.** Demographics.

| Characteristic |  |
|---|---|
| Age |  |
| Mean (SD) | 47.2 (10.30) |
| Median (IQR) | 45.5 (41.0, 55.0) |
| Race, n (%) |  |
| Black | 13 (50.0%) |
| White | 13 (50.0%) |
| Sex, n (%) |  |
| Female | 15 (57.7%) |
| Male | 11 (42.3%) |
| FTND Total Score |  |
| Mean (SD) | 5.1 (2.01) |
| Median (IQR) | 5.0 (4.0, 7.0) |
| Average cigarettes per day |  |
| Mean (SD) | 14.4 (6.31) |
| Median (IQR) | 12.0 (10.0, 20.0) |
| Years smoking |  |
| Mean (SD) | 23.3 (12.61) |
| Median (IQR) | 21.5 (14.0, 30.0) |

*Note.* N = 26. FTND = Fagerström Test for Nicotine Dependence

Overall, the devices significantly differed in trends over time (p<0.001), highlighted by the high values seen in the corded Bedfont iCOquit in the later time periods. For the remaining models, only the Vitalograph BreathCO readings were considered. **Error! Reference source not found**.2B shows difference in CO by sleep condition. Surprisingly, this difference is significant, p=0.0291, despite little difference being visible apart from a higher starting CO for the sleep extension condition. Figure 2C illustrates the significantly greater values of breath CO among the White participants (p<0.001), indicating higher baseline cigarettes per day. Figure 2D illustrates the statistically significant difference between sexes on breath CO patterns (p=0.005), though the difference is not visually prominent. The Kaplan-Meier curve of CO measured by the Vitalograph BreathCO (Figure 3) shows that 50% of participants reached CO ≤3 ppm before 21 hours, and all participants reached that level by 42 hours. Interestingly, there was a sharp increase in the prevalence of CO ≤3 ppm at the 12-hour timepoint, which may be due to the timing of measurements after the first night’s sleep.

Examining efficiency, we see from Tables 2 to 4 that both the corded Bedfont iCOquit and Bluetooth iCOquit values of 3 ppm best predicted a Vitalograph BreathCO reading of ≤3 ppm, but the piCO+ was most predictive of the Vitalograph BreathCO at 5 ppm. From Figures 1 and 2, we see that piCO+ had consistently greater average readings than the Vitalograph BreathCO, while the Bluetooth iCOquit had average readings similar to the Vitalograph BreathCO. Both devices had similar efficiencies (86%). While the corded iCOquit had the same cutoff as the Vitalograph BreathCO, it had the lowest predictive measures (efficiency, Youden’s J, etc.) with noted variability in measurements across time.

**Table 2.** Results of ROC Analyses.

| Cutoff | Sensitivity | Specificity | PPV | NPV | Youden’s J | Efficiency | AUC |
|---|---|---|---|---|---|---|---|
| piCO+ predicting Vitalograph BreathCO ≤ 3 |  |  |  |  |  |  |  |
| 0 | 21.4 | 95.0 | 84.0 | 49.4 | 0.164 | 0.543 | 0.582 |
| 1 | 24.2 | 94.4 | 84.3 | 50.1 | 0.186 | 0.556 | 0.593 |
| 2 | 41.8 | 93.6 | 89.0 | 56.5 | 0.354 | 0.650 | 0.677 |
| 3 | 64.5 | 92.0 | 90.9 | 67.6 | 0.565 | 0.768 | 0.783 |
| 4 | 80.1 | 87.0 | 88.4 | 77.9 | 0.671 | 0.832 | 0.835 |
| **5** | **89.3** | **81.2** | **85.5** | **86.0** | **0.705** | **0.857** | **0.852** |
| 6 | 94.2 | 69.8 | 79.4 | 90.7 | 0.640 | 0.833 | 0.820 |
| 7 | 97.0 | 60.2 | 75.1 | 94.2 | 0.572 | 0.805 | 0.786 |
| Corded iCOquit predicting Vitalograph BreathCO ≤ 3 |  |  |  |  |  |  |  |
| 0 | 17.7 | 96.1 | 85.1 | 48.2 | 0.138 | 0.528 | 0.569 |
| 1 | 46.4 | 92.8 | 89.0 | 58.0 | 0.392 | 0.672 | 0.696 |
| 2 | 63.2 | 87.7 | 86.6 | 65.5 | 0.508 | 0.741 | 0.754 |
| **3** | **76.3** | **76.9** | **80.6** | **72.1** | **0.532** | **0.766** | **0.766** |
| 4 | 81.8 | 67.6 | 76.0 | 74.8 | 0.494 | 0.754 | 0.747 |
| 5 | 85.4 | 60.4 | 73.0 | 76.7 | 0.458 | 0.742 | 0.729 |
| 6 | 87.3 | 55.6 | 71.2 | 77.7 | 0.429 | 0.731 | 0.714 |
| 7 | 87.6 | 51.7 | 69.4 | 76.8 | 0.392 | 0.715 | 0.696 |
| Bluetooth iCOquit predicting Vitalograph BreathCO ≤ 3 |  |  |  |  |  |  |  |
| 0 | 22.7 | 96.5 | 87.7 | 53.0 | 0.192 | 0.557 | 0.596 |
| 1 | 55.5 | 94.0 | 91.0 | 65.6 | 0.494 | 0.727 | 0.747 |
| 2 | 75.9 | 88.4 | 87.9 | 76.9 | 0.644 | 0.815 | 0.822 |
| **3** | **90.5** | **79.9** | **83.3** | **88.3** | **0.704** | **0.857** | **0.852** |
| 4 | 95.5 | 67.3 | 76.4 | 93.1 | 0.628 | 0.829 | 0.814 |
| 5 | 98.2 | 54.8 | 70.6 | 96.5 | 0.530 | 0.787 | 0.765 |
| 6 | 100.0 | 46.2 | 67.3 | 100.0 | 0.462 | 0.759 | 0.731 |
| 7 | 0.0 | 100.0 | - | 47.5 | 0.000 | 0.448 | 0.686 |

*Note.* Bolded rows indicate the highest efficiency for that device. PPV = positive predictive value. NPV = negative predictive value. AUC = area under the curve.

**Figure 1.**
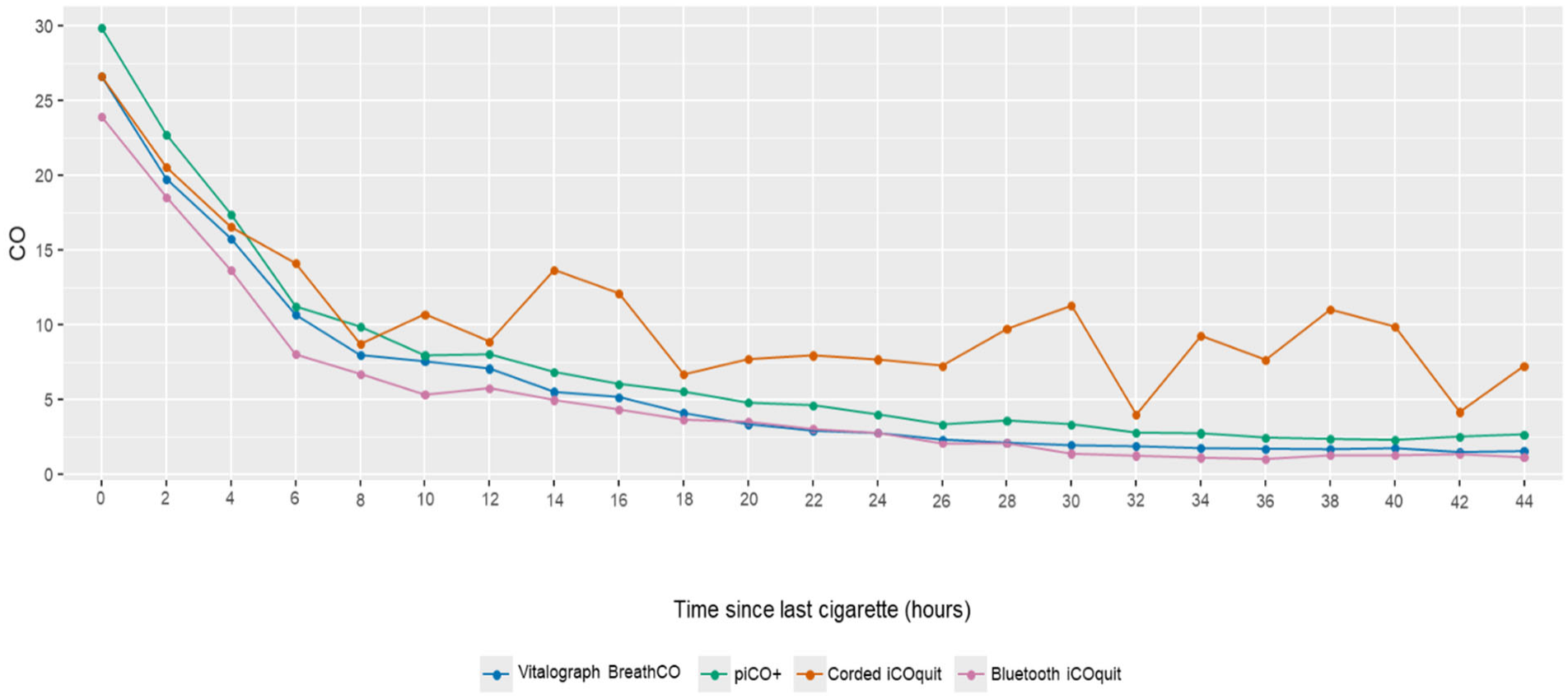
CO raw means over time by instrument

**Figure 2.**
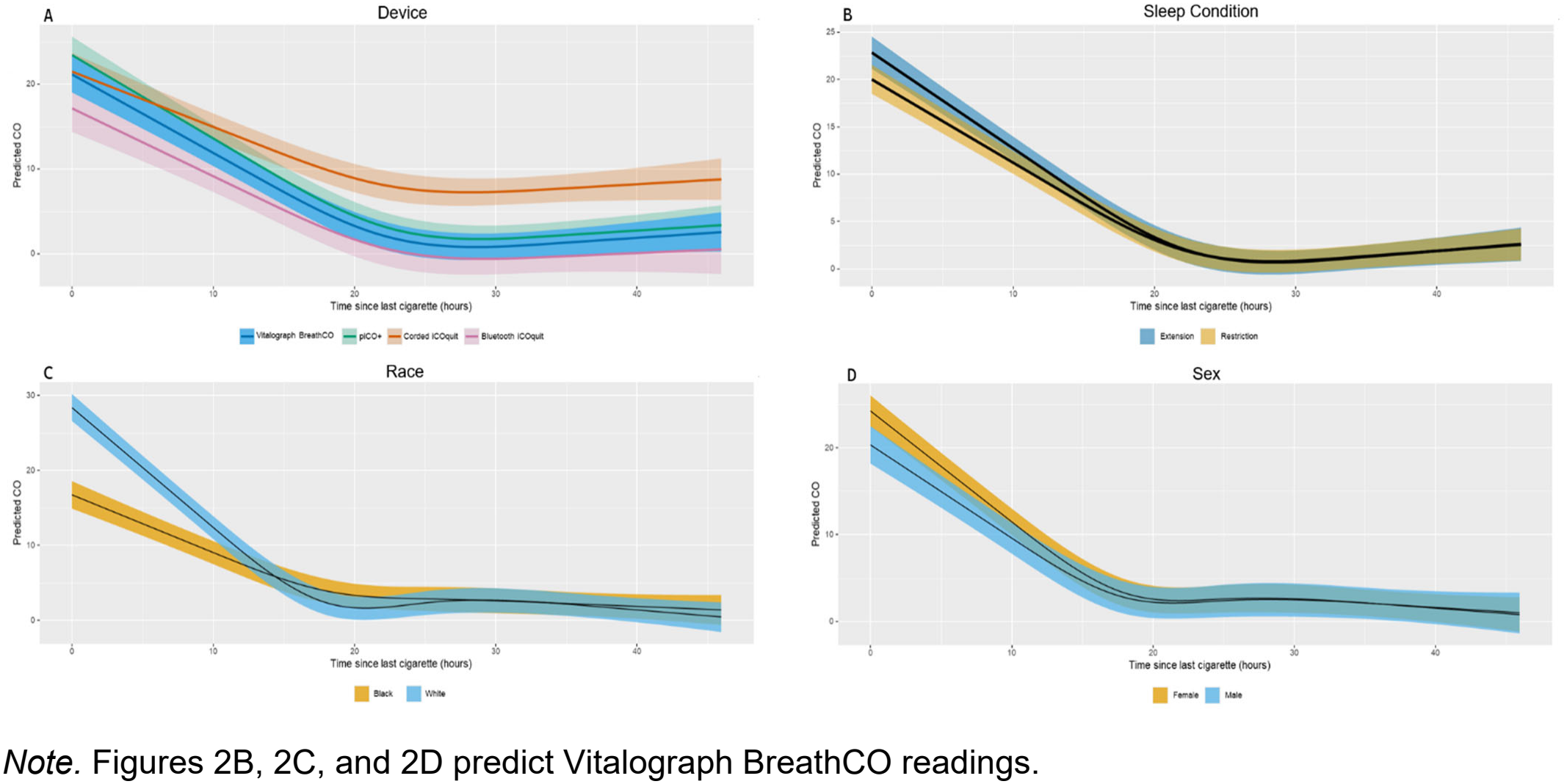
Predicted CO over time by device, sleep condition, race, and sex

**Figure 3.**
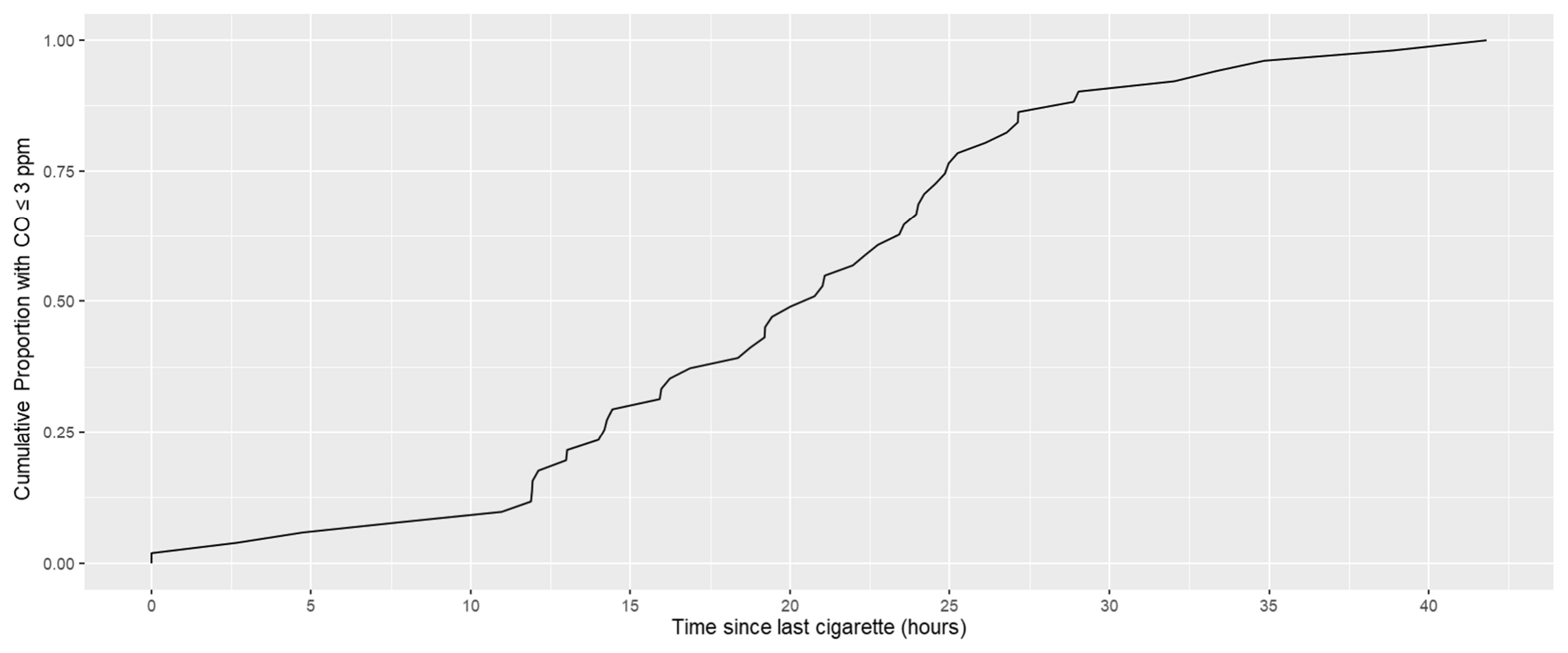
Time for participants to achieve CO ≤ 3 ppm from Vitalograph BreathCO

## 4 DISCUSSION

These results are consistent with previous findings that readings from different models of breath CO monitors should not be used interchangeably (Tonkin et al., 2023; Tuck et al., 2021). When using breath CO reading cutoffs for determining abstinence, researchers should consider a value specific to each device. For instance, in this study, a piCO+ reading of 5 ppm resulted in the highest efficiency for that device and matched a Vitalograph BreathCO reading of ≤3 ppm. A reading of 3 ppm did provide the highest efficiency for the Bluetooth and corded versions of the iCOquit. However, though the best value matched in those cases, the maximum efficiency of the corded iCOquit was very low at 0.77. Additionally, the change in mean CO readings over time from the corded iCOquit did not follow a smooth decline, especially relative to the other devices. While we did not measure these devices against CO canisters with standardized concentrations, previous research demonstrated consistently low readings from the bluetooth iCOquit and piCO+ whereas the Vitalograph BreathCO matched the 5 ppm canister and showed 11 ppm on a 10 ppm canister (Tonkin et al., 2023). Overall, CO readings are particular to the device used, so it is essential to use the same device through the duration of a study.

This study illustrated patterns in breath CO dissipation over the course of 44 hours. The pattern of mean CO concentration over time differed between devices. Most prominently, after the initial drop, the corded iCOquit recorded higher average values than the other devices. Surprisingly, there was also a significant difference between trends based on sleep condition. Likely this significance is due, in part, to the amount of data collected (n=26 participants with up to 21 measurements over two days and two sessions giving a total of 851 observations in each model). Breath CO patterns also differed based on race and sex, with White and male participants having higher average readings than Black and female participants, respectively.

These differences primarily appeared at the initial readings, which reflects the known pattern of higher cigarettes per day by men (Wright et al., 2026) and White people who smoke (Nguyen-Grozavu et al., 2020). The race- and sex-based differences on breath CO were not prominent 24 hours after the participants’ last cigarettes, indicating that breath CO cutoffs for determining smoking abstinence do not need to be dependent on those characteristics.

These results should be considered in the context of the study’s limitations. Though each participant’s last cigarette was smoked at a standardized time, and participants reported they smoked at least five cigarettes per day on average to be eligible, there was still variability in how recently and how heavily participants had smoked before beginning study procedures.

Furthermore, while we measured CO every two hours during waking hours, participants slept for at least four hours, during which time CO was not measured. This likely accounts for some of the sudden drops in CO measurement. However, given that the timing of sleep opportunities varied between participants based on their normal sleep schedule, we were able to capture timepoints across all two-hour bins of measurement. All participants were also either White or Black, limiting the ability to generalize to other racial/ethnic groups. Finally, the devices were not compared against known CO concentrations, although previous research has shown that all devices have some variability in measurement (Tonkin et al., 2023).

Strengths of this study include the ability to continuously monitor participants and determine the time course of breath CO dissipation over two nights of observed nicotine abstinence. Moreover, we examined how the pattern of CO dissipation differed between the four most used breath CO devices, including the two iCOquit devices that are increasingly being used in remote clinical trials. Finally, while we only enrolled 26 participants, relatively equal numbers of men and women and White and Black participants with multiple data collection timepoints provided robust measurement of CO values across time, lending confidence in the findings.

Overall, individual participant characteristics did not have a notable impact on breath CO after a day of smoking abstinence, but the readings varied based on the device used. The Vitalograph BreathCO was used as the gold standard, and ROC analysis found the piCO+ and Bluetooth iCOquit, both from Bedfont, to have maximum efficiencies of 0.86 for detecting a Vitalograph BreathCO reading of ≤3 ppm. However, the piCO+ cutoff with maximum efficiency was 5 ppm, rather than 3 ppm. The corded version of the Bedfont iCOquit had a maximum efficiency when the cutoff was 3 ppm, but that efficiency was only 0.77, indicating more variability in measurement. Thus, researchers should note that different models of breath CO devices have different optimal cutoff values for determining smoking abstinence, and some devices produce more consistent values than others.

Funding for this project was provided to MPIs KLC/KLG by NIH R01DA046096. Conflicts of Interest:

## Data Availability

All data produced in the present study are available upon reasonable request to the authors.

## References

Agaku, I. (2026). Tobacco Product Use among U.S. Adults, 2023–2024. NEJM Evidence, 5(4), EVIDpha2500339. 10.1056/EVIDpha2500339

Arrazola, R. A. (2025). Notes from the Field: Tobacco Product Use Among Adults — United States, 2017–2023. MMWR. Morbidity and Mortality Weekly Report, 74. 10.15585/mmwr.mm7407a3

Benowitz, N. L., Bernert, J. T., Foulds, J., Hecht, S. S., Jacob, P., Jarvis, M. J., Joseph, A., Oncken, C., & Piper, M. E. (2020a). Biochemical Verification of Tobacco Use and Abstinence: 2019 Update. Nicotine & Tobacco Research : Official Journal of the Society for Research on Nicotine and Tobacco, 22(7), 1086–1097. 10.1093/ntr/ntz132

Benowitz, N. L., Bernert, J. T., Foulds, J., Hecht, S. S., Jacob, P., Jarvis, M. J., Joseph, A., Oncken, C., & Piper, M. E. (2020b). Biochemical Verification of Tobacco Use and Abstinence: 2019 Update. Nicotine & Tobacco Research: Official Journal of the Society for Research on Nicotine and Tobacco, 22(7), 1086–1097. 10.1093/ntr/ntz132

Cropsey, K. L., Eldridge, G. D., Weaver, M. F., Villalobos, G. C., & Stitzer, M. L. (2006). Expired carbon monoxide levels in self-reported smokers and nonsmokers in prison. Nicotine & Tobacco Research: Official Journal of the Society for Research on Nicotine and Tobacco, 8(5), 653–659. 10.1080/14622200600789684

Cropsey, K. L., Trent, L. R., Clark, C. B., Stevens, E. N., Lahti, A. C., & Hendricks, P. S. (2014). How low should you go? Determining the optimal cutoff for exhaled carbon monoxide to confirm smoking abstinence when using cotinine as reference. Nicotine & Tobacco Research: Official Journal of the Society for Research on Nicotine and Tobacco, 16(10), 1348–1355. 10.1093/ntr/ntu085

Cummings, K. M. (2016). Smoking Isn’t Cool Anymore: The Success and Continuing Challenge of Public Health Efforts to Reduce Smoking. Journal of Public Health Management and Practice : JPHMP, 22(1), 5–8. 10.1097/PHH.0000000000000360

Dahne, J., Wahlquist, A. E., McClure, E. A., Natale, N., Carpenter, M. J., & Tomko, R. L. (2024). Remote Carbon Monoxide Capture via REDCap: Evaluation of an Integrated Mobile Application. Nicotine & Tobacco Research: Official Journal of the Society for Research on Nicotine and Tobacco, 26(6), 696–703. 10.1093/ntr/ntad230

Doll, R., Peto, R., Boreham, J., & Sutherland, I. (2004). Mortality in relation to smoking: 50 years’ observations on male British doctors. BMJ (Clinical Research Ed.), 328(7455), 1519. 10.1136/bmj.38142.554479.AE

Hastie, T., Tibshrani, R., & Friedman, J. (2009). Elements of Statistical Learning: Data Mining, Inference, and Prediction (2nd ed.). https://hastie.su.domains/ElemStatLearn/

Nguyen-Grozavu, F. T., Pierce, J. P., Sakuma, K.-L. K., Leas, E. C., McMenamin, S. B., Kealey, S., Benmarhnia, T., Emery, S. L., White, M. M., Fagan, P., & Trinidad, D. R. (2020). Widening disparities in cigarette smoking by race/ethnicity across education level in the United States. Preventive Medicine, 139, 106220. 10.1016/j.ypmed.2020.106220

Ramani, V. K., Mhaske, M., & Naik, R. (2023). Assessment of Carbon Monoxide in Exhaled Breath using the Smokerlyzer Handheld Machine: A Cross-Sectional Study. Tobacco Use Insights, 16, 1179173X231184129. 10.1177/1179173X231184129

SRNT Subcommittee on Biochemical Verification. (2002). Biochemical verification of tobacco use and cessation. Nicotine & Tobacco Research, 4(2), 149–159. 10.1080/14622200210123581

Thrul, J., Howe, C. L., Devkota, J., Alexander, A., Allen, A. M., Businelle, M. S., Hébert, E. T., Heffner, J. L., Kendzor, D. E., Ra, C. K., & Gordon, J. S. (2023). A Scoping Review and Meta-analysis of the Use of Remote Biochemical Verification Methods of Smoking Status in Tobacco Research. Nicotine & Tobacco Research: Official Journal of the Society for Research on Nicotine and Tobacco, 25(8), 1413–1423. 10.1093/ntr/ntac271

Tonkin, S., Kezbers, K. M., Noble, B., Cropsey, K., Kendzor, D. E., Oliver, J., & Businelle, M. (2023). Comparison of the Bluetooth iCOquit, piCO, and Vitalograph for the assessment of breath carbon monoxide among adults initiating smoking cessation and standardized canisters. Drug and Alcohol Dependence, 250, 110902. 10.1016/j.drugalcdep.2023.110902

Tuck, B. M., Karelitz, J. L., Tomko, R. L., Dahne, J., Cato, P., & McClure, E. A. (2021). Mobile, Remote, and Individual Focused: Comparing Breath Carbon Monoxide Readings and Abstinence Between Smartphone-Enabled and Stand-Alone Monitors. Nicotine & Tobacco Research: Official Journal of the Society for Research on Nicotine and Tobacco, 23(4), 741–747. 10.1093/ntr/ntaa203

VanFrank, B., Malarcher, A., Cornelius, M. E., Schecter, A., Jamal, A., & Tynan, M. (2024). Adult Smoking Cessation—United States, 2022. MMWR. Morbidity and Mortality Weekly Report, 73(29), 633–641. 10.15585/mmwr.mm7329a1

Wright, L., Kock, L., Tattan-Birch, H., & Bann, D. (2026). Cigarette smoking across life from 1946 to 2018: Harmonisation of four British birth cohort studies. Addiction, 121(2), 440–447. 10.1111/add.70204

